# The Impact of HPV DNA/p16 in Laryngeal/Hypopharyngeal Cancer: a Systematic Review and Meta-analysis

**DOI:** 10.1101/2021.04.10.21255247

**Authors:** Sarah Van der Elst, Daniel P. Russo, Derek Mumaw, Michael Wotman, Tristan Tham

## Abstract

**Background:** This meta-analysis seeks to investigate the association between HPV and p16 status with overall survival in laryngeal and hypopharyngeal carcinoma.

**Methods:** Medline, Scopus, EMBASE, and the Cochrane Library were used to identify studies for inclusion. Abstracts that discussed HPV/p16 status and prognosis in laryngeal or hypopharyngeal carcinoma were included. Next, full-text articles were screened and included based upon a checklist established *a priori*. Pooled hazard ratios for overall survival were generated using a random effects model. RevMan 5.3, Meta Essentials, and OpenMeta[Analyst] were used for statistical analysis.

**Results:** Thirteen studies published between 2014 and 2019 with sample sizes ranging from 31 to 9,656 were selected for inclusion in this meta-analysis. The pooled data demonstrated that p16 status was not significantly associated with OS in either laryngeal or hypopharyngeal carcinoma with HRs of 1.03 (95% CI: 0.73–1.45; p = 0.88) and 1.02 (95% CI: 0.55–1.86; p = 0.96), respectively. The pooled data showed that HPV status was predictive of OS in laryngeal cancer with 0.63 (95% CI: 0.41–0.97; p = 0.03).

**Conclusions:** Our results suggest that p16-positivity does not provide a survival benefit in LC and HPC. This is in contrast to studies in the oropharynx, where p16 status is a standard proxy for HPV infection and HPV infection is associated with improved prognosis.

## Introduction

Head-and-neck cancer (HNC) represents malignancies that occur in the nasopharynx, oropharynx, oral cavity, and larynx and are some of the most common malignancies worldwide (1). While a substantial number of cases has traditionally been attributed to alcohol and tobacco use, Human Papillomavirus (HPV) infection has been implicated in squamous cell carcinoma of the head and neck (HNSCC) within a variety of the aforementioned locations (2). High-risk HPV DNA, most commonly HPV-16, encodes E6 and E7 oncoproteins, which are theorized to deactivate tumor suppressors p53 and pRb, respectively, to induce tumorigenesis. By this mechanism, cell cycle protein p16 is believed to rise in active HPV infection.

Tumors are defined as HPV-positive when both HPV DNA and direct transcripts of E6/E7 mRNA are positive (3). For oropharyngeal squamous cell carcinoma (OPSCC), p16 positivity serves as a proxy for HPV DNA because of the consistent positive correlation between the two markers (4-6). By contrast, this relationship has not yet been established in the larynx and hypopharynx, where other non-viral mechanisms may explain p16 positivity (7-9). Therefore, in LC and HPC, laboratory evidence of both HPV DNA and E6/E7 mRNA transcripts remains the standard for determination of tumor positivity for HPV.

The prognostic effect of HPV and p16 status in OPSCC is well established (10). Indeed, the AJCC 8^th^ edition in staging rules has been modified to classify this cancer separately because of the superior prognostic outcomes (11). Specifically, this edition distinguishes OPSCC associated with high-risk HPV strains from OPSCC not associated with such strains (11). In contrast to the observed correlation within the oropharynx, the same connection for laryngeal squamous cell carcinoma (LC) and hypopharyngeal squamous cell carcinoma (HPC) has not yet been established (12, 13).

Similarly, the prognostic effect of p16 positivity in LC and HPC has been investigated in the literature with conflicting results. For example, some research has indicated that p16 expression in LC is correlated with increased survival (14, 15). However, in direct contradiction of these conclusions, others, such as Dahm and et al. at the Medical University of Austria, have maintained that p16 tumor positivity does not impact survival time for these patients, instead describing smoking status and tumor staging as significant prognostic indicators (16-18).

Although studies have investigated the association of both HPV (HPV DNA and E6/E7 mRNA transcripts) and p16 status with outcomes in LC and HPC, no study has to our knowledge synthesized the literature to compile the evidence on these relationships directly. In this study, we synthesized the available literature and conducted a meta-analysis to (1) clarify the current ambiguity in the prognostic role of p16-positivity positivity in laryngeal and hypo-pharyngeal carcinoma and (2) investigate the prognostic impact of HPV-positivity within these subsites.

## Materials and Methods

### Study Design

This study was designed in accordance with several guidelines and checklists. The search followed the guidelines described in the chapter on searching of the *Cochrane Handbook for DTA Reviews* (19). To further clarify the protocols in this review, we utilized the Preferred Reporting Items for Systematic Reviews and Meta-Analyses (PRISMA) statement guidelines (20). The study was designed *a priori* and registered in the PROSPERO online database (21).

### Search Strategy

On February 2, 2019, we searched PubMed, Scopase, Embase, and the Cochrane Library from their inception to the present. The following terms were used for the purposes of this search: laryngeal carcinoma, hypopharyngeal carcinoma, and human papillomavirus. Any variation of the aforementioned terms was also included during the initial search. The full strategy is described in the supplemental materials (S1).

### Article Selection

Selection of studies was performed by two authors (TT & DM) in two phases. In the first phase, abstracts were screened to establish relevance to the research question. Any abstract that discussed HPV/p16 status and prognosis in laryngeal or hypopharyngeal carcinoma was included for further evaluation in the second phase of screening. In the second phase, full-text articles were evaluated by the aforementioned authors based upon inclusion and exclusion criteria that were determined a priori. The inclusion and exclusion criteria are provided in Table 1. The list of full-text articles retrieved for further analysis is outlined in the supplemental materials (S2).

**Table 1.**
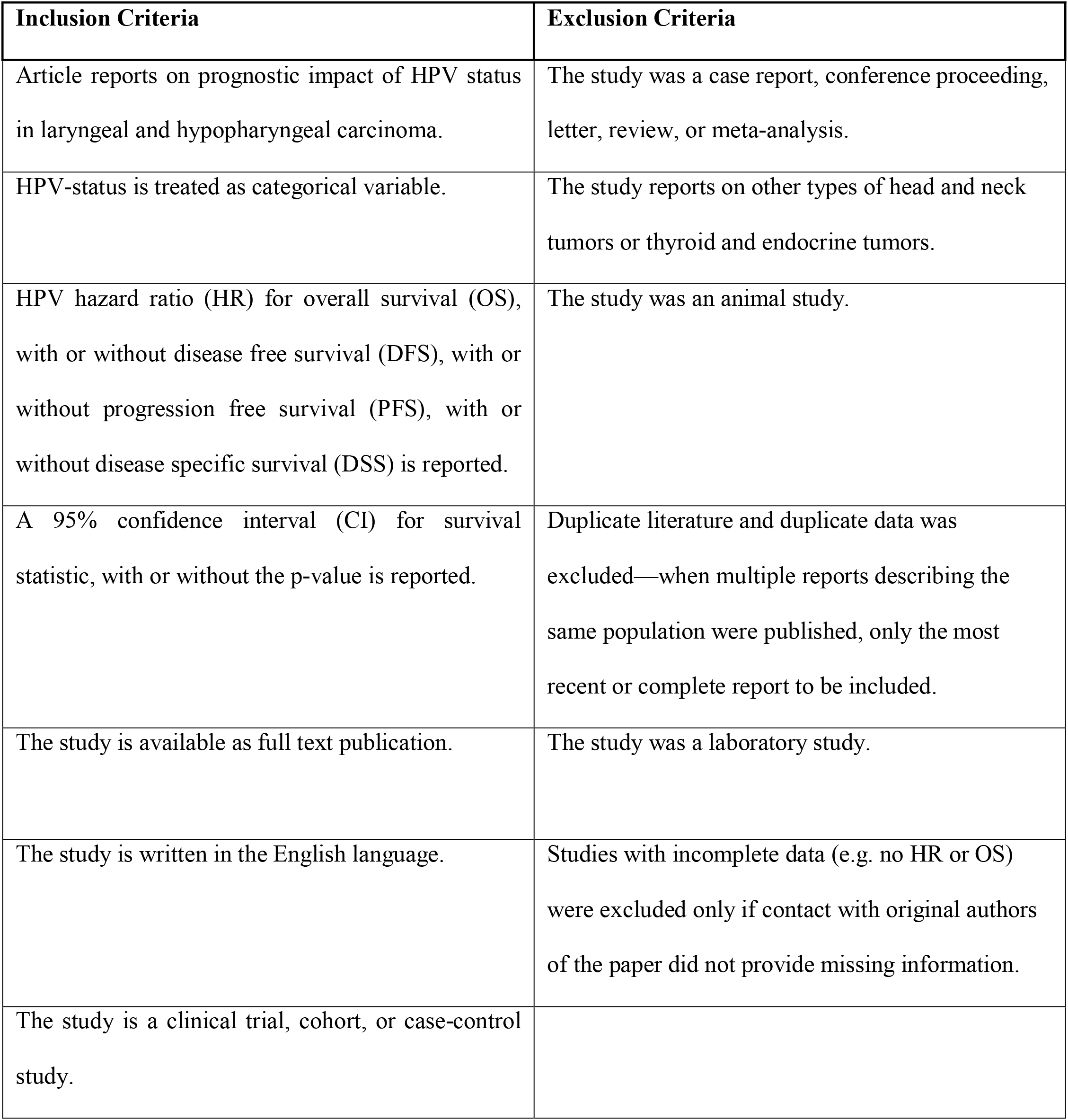
Inclusion and exclusion criteria for full-text studies.

This search strategy yielded 1,670 total studies after initial article selection. Following phase one screening, 94 abstracts were eligible for full-text retrieval. After the second phase of screening full-text articles, 14 articles were included in this study. The PRISMA flow sheet for this method is included in Figure 1. An outline of the articles excluded from this study with reasons can be found in the supplemental materials (S3).

**Figure 1.**
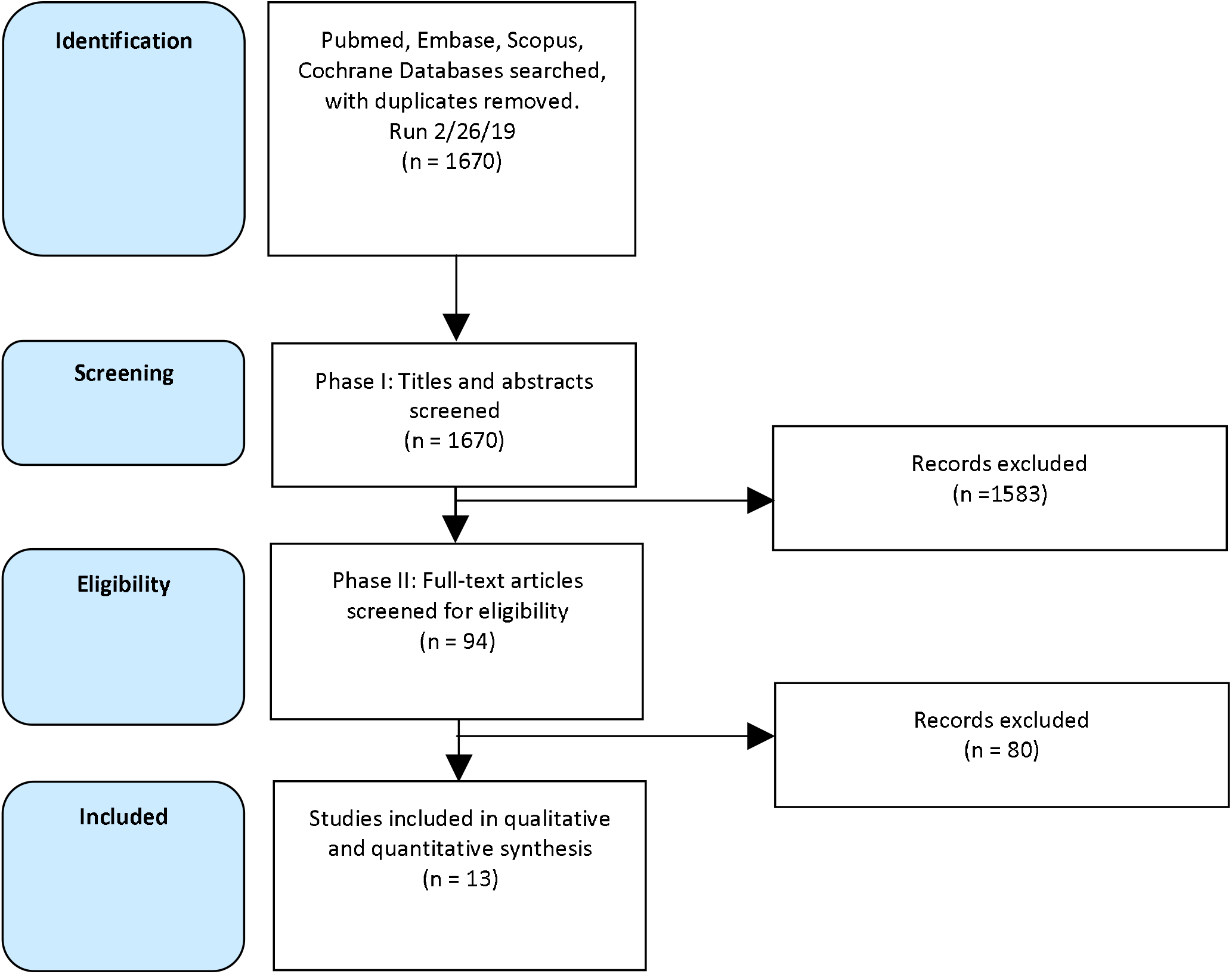
**Preferred Reporting Items for Systematic Reviews and Meta-Analysis (PRISMA) Flowchart**

### Quality Assessment

Two authors (TT & DM) assessed the risk for bias in the included studies using the Quality In Prognosis Studies (QUIPS) Tool (22). This quality-assessment tool determines the risk of bias based upon six domains: (1) Study Participation; (2) Study Attrition; (3) Prognostic Factor Measurement; (4) Outcome Measurement; (5) Study Confounding; and (6) Statistical Analysis and Reporting. Through the scoring system, each of the six domains carries a score of low, moderate, and high risk for bias. The composite score of all six domains describes the risk for bias in the entire study. The average composite score across the domains was then calculated and can be found in the supplemental materials (S4).

### Data Extraction

Two of the authors (TT & DM) reviewed all of the full-text articles to extract the data. Several data points were collected during this stage: (1) first author’s name; (2) publication year; (3) sample size; (4) country/state (if USA) of the study population; (5) patient population (e.g., hospital); (6) study design; (7) tumor site; (8) follow-up period; (9) average age of the patient population; (10) tumor staging; (11) HPV prevalence; (12) p16 prevalence; (13) HPV detection methodology; (14) outcome measure; (15) treatment type; (16) HR for OS (with 95% CI); and (17) HR for DFS/RFS, if applicable (with 95% CI). In studies that only reported the KM analysis without the hazard ratio, we used the methods described by Parmar and Tierney to reconstruct the data (23, 24).

### Statistical Analysis

The primary statistic for this study was the logarithm of the HR (Log[HR]) with standard error (SE). The HR with 95% CI, which was obtained during data extraction, was utilized to calculate these statistics. Estimates of the Log[HR] were weighted and pooled using the generic inverse-variance method. The random effects model described by the DerSermonian and Laird methods was chosen for analyses as a conservative approach given the risk for heterogeneity. Heterogeneity was further assessed using the Cochrane’s Q and Higgins I^2^. Significant heterogeneity was defined as a Cochrane’s Q of p<0.1 and an I^2^> 50%. Exploration of heterogeneity was attempted if detected, but if no source was found, we elected not to pool the individual HRs to a pooled HR (25). All analyses were completed using the RevMan 5.3 analysis software (Cochrane Collaboration, Copenhagen, Denmark) (26). Statistical tests were two-sided. Statistical significance was defined as a p-value <0.05.

### Publication Bias

To assess publication bias, Begg’s Funnel Plot and the Egger’s Bias Indicator test were used. We utilized Meta-Essentials (ERASMUS Research Institute, Rotterdam, Netherlands) to perform tests of publication bias (27). The results of these tests can be found in the supplemental materials (S5, S7).

## Results

### Study Characteristics

Thirteen studies published between 2014 and 2019 with sample sizes ranging from 45 to 9,656 were selected for inclusion in this meta-analysis (7, 15, 28-39). The characteristics of the studies can be found in Table 2. Three studies were from the USA, four were from China, one was from Spain, one was from Sweden, one was from Australia, one was from Brazil, one was from South Korea, and one was from Singapore. Eleven of the studies were retrospective cohort studies, one was a retrospective chart review, and one was a registry database study. Ten studies reported data on LC, and four studies reported data on HPC.

**Table 2.** Study Characteristics (7, 15, 28-38)

| Author | Year | Country | Study Design | Age (years) | Site | Total (n) | HPV DNA Prevalence | P16 Prevalence | HPV Detection Method | p16 Detection Method | Outcome |
| --- | --- | --- | --- | --- | --- | --- | --- | --- | --- | --- | --- |
| Wang | 2015 | China | RCR | 59 | L | 318 | 13.2 | 3 | HPV DNA PCR | IHC: >70% Staining | OS, RFS, DMFS |
| Tong | 2018 | China | RCS | 58.2 | L | 211 | 62.5 | 54 | HPV PCR and HPV DNA ISH | IHC: >75% Staining | OS |
| Sanchez-Barrueco | 2019 | Spain | RCS | NR | L | 123 | 22.76 | 39.02 | HPV DNA PCR | IHC: >70% Staining | OS, DFS |
| Young | 2015 | Australia | RCS | 66 | L | 307 | 5.1 | 6.5 | HPV RNA ISH | IHC: >30% Staining and Staining Intensity | OS, FFS |
| Yang | 2016 | China | RCS | NR | L | 196 | 16.8 | NR | HPV DNA qPCR | IHC: >70% Staining | OS, PFS |
| Wendt | 2014 | Sweden | RCS | 68 | HP | 109 | 6 | 16 | HPV DNA PCR | IHC: 3-Tier Scale | OS, DFS |
| Lopez | 2014 | Brazil | RCS | NR | L | 142 | 12.7 | NR | HPV DNA PCR & HPV E6/E7 ELISA | N/A | OS |
| Lam | 2018 | China | RCSS | 68.6 | L | 85 | 1.2 | 12.9 | HPV 16 & E5 DNA | IHC: focal (<25%) vs. diffuse (>25%) | OS, DSS |
| Li | 2018 | USA | RCS | 63.1 | LHP | 9656 | 13.12 | NR | NCDB Detection Method | N/A | OS |
| Lee | 2018 | South Korea | RCS | 63 | HP | 45 | NR | 24.4 | N/A | IHC: Graded staining | OS, PFS |
| Ang | 2014 | Singapore | RCS | 65.4 | HP | 75 | NR | 6.67 | HPV DNA ISH | IHC: >70% Staining | OS |
| Fakhry | 2017 | USA | RCS | 58 | L | 243 | 5 | 13 | HPV DNA ISH | IHC: 70% Staining | OS |
| Hernandez | 2014 | USA | Registry Study | NR | L | 178 | 21 | NR | HPV DNA PCR | N/A | OS |

RCR, retrospective chart review; RCS, retrospective cohort study; RCSS, retrospective cross sectional study; L, larynx; HP, hypopharynx; LHP, larynx and hypopharynx; NCDB, National Cancer Database; OS, overall survival; DFS, disease-free survival; RFS, relapse-free survival; DMFS, distant metastasis-free survival; DSS, disease specific survival; PFS, progression-free survival; FSS, failure-free survival; S, surgery; RT, radiotherapy; CT, chemotherapy; NR, not reported; ISH, *in situ* hybridization; IHC, immunohistochemistry; PCR, polymerase chain reaction; qPCR, quantitative PCR.

In terms of outcome, six studies reported overall survival (OS); two OS and progression-free survival (PFS); two OS and disease-free survival (DFS); one OS and failure-free survival (FSS); one OS and disease-specific survival (DSS); and one OS, DMFS, and relapse-free survival (RFS). HR was reported for twelve studies and was extrapolated for one study from a KM curve. Summaries of survival outcomes are described in Table 3.

**Table 3.** Summary of Survival Outcomes (7, 15, 28-38)

| **Larynx** |  |  |  |
| --- | --- | --- | --- |
| Author | Comparator Groups | OS |  |
|  |  | *HR (95% CI)* | *p* value |
| Wang | HPV+ vs. HPV-/p16+ vs. p16- | **HPV:** 0.34 (0.12-.97) | 0.045 |
|  |  | **p16:** 1.83 (0.57-5.86) | 0.31 |
| Tong | HPV+ vs. HPV- | **HPV:** 0.395 (0.185-0.843) | 0.016 |
|  |  | **p16:** 0.837 (0.375-1.868) | 0.664 |
| Barrueco<sup>1</sup> | P16+ vs. p16- | 1.32 (0.64-2.73) | 0.43 |
| Young | P16+ vs. p16- | 0.74 (0.32-1.69) | 0.48 |
| Fakhry | HPV+ vs. HPV-/p16+ vs. p16- | 1.15 (0.54-2.46) | 0.714 |
| Lopez | HPV16 DNA+ vs. HPV16 DNA- | 0.76 (0.09-6.23) | 0.796 |
| Lam | p16+ vs. p16- | 0.7(0.25-1.97) | 0.5 |
| Li | HPV+ vs. HPV- | 0.71 (0.59-0.85) | NR |
| Hernandez | HPV+ vs. HPV- | 1.28 (0.66-2.51) | NR |
| Yang | HPV+ vs. HPV- | 0.18 (0.057, 0.58) | 0.004 |
| **Hypopharynx** |  |  |  |
| Author | Comparator Groups | OS |  |
|  |  | *HR (95% CI)* | *p* value |
| Wendt | HPV+ vs. HPV-/p16+ vs. p16- | 0.16 (0.034-0.7) | 0.015 |
|  |  | **p16:** 1.31 (0.61-2.80) | 0.49 |
|  |  | **HPV:** 0.10 (0.012-0.81) | 0.031 |
| Li | HPV+ vs. HPV- | 0.59 (0.45-0.77) | NR |
| Lee | p16+ vs. p16- | 0.601 (0.153-2.366) | 0.466 |
| Ang | p16+ vs. p16-/ Cyclin D1+ vs. Cyclin D1- | 0.73 (0.17-3.12) | 0.671 |

OS, overall survival; HR, hazard ratio; CI, confidence interval; NR, not reported.
<sup>1</sup>Data was reconstructed from the reported Kaplan-Meier curve to obtain reported HR.

Publication bias assessment was only assessed for p16 status and OS in LC and for HPV status and OS in the HPC data groups as there were inadequate numbers of studies in all other groups to perform more advanced statistical tests. Funnel plots are included in the supplementary materials.

### P16 status and OS in hypopharyngeal cancer

The data from three studies was synthesized into the meta-analysis on p16 and OS in patients with HPC. The pooled data showed that there was not a significant association between p16 status and OS in HPC with a HR of 1.02 (95% CI: 0.55, 1.86; p-value = 0.96) (Figure 2). There was little heterogeneity with an I^2^ of 22% and a p-value of 0.28. Because of the smaller number of studies included in this analysis, subgroup analysis was not performed and publication bias was not assessed.

**Figure 2.**
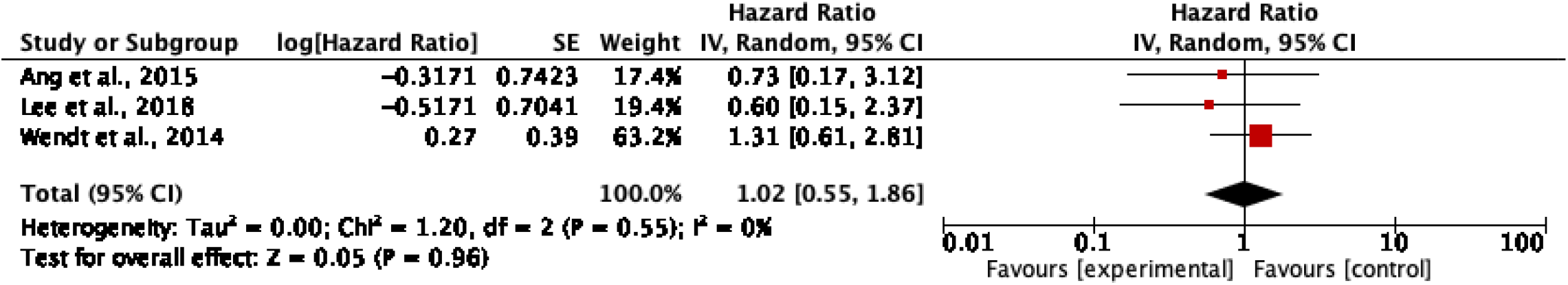
Association between p16 status and OS in hypopharyngeal carcinoma Values on the right of the forest plot indicate reduced survival with p16 positivity. Meanwhile, values on the left of the plot indicate reduced survival with p-16 negativity. The diamond represents the pooled result for all studies included in the analysis.

### P16 status and OS in laryngeal cancer

Data from six studies was collected for the meta-analysis on p16 and OS in patients with LC. A significant association between p16 and OS was not found with a HR of 1.03 (95% CI: 0.73, 1.45; p-value = 0.88) (Figure 3). No heterogeneity was found with an I^2^ of 0% and a p-value of 0.71. Begg’s funnel plot for HR of OS did not suggest publication bias. The p value for Egger’s test also indicated that there was no publication bias for OS (p = 0.973) (See Supplemental Materials).

**Figure 3.**
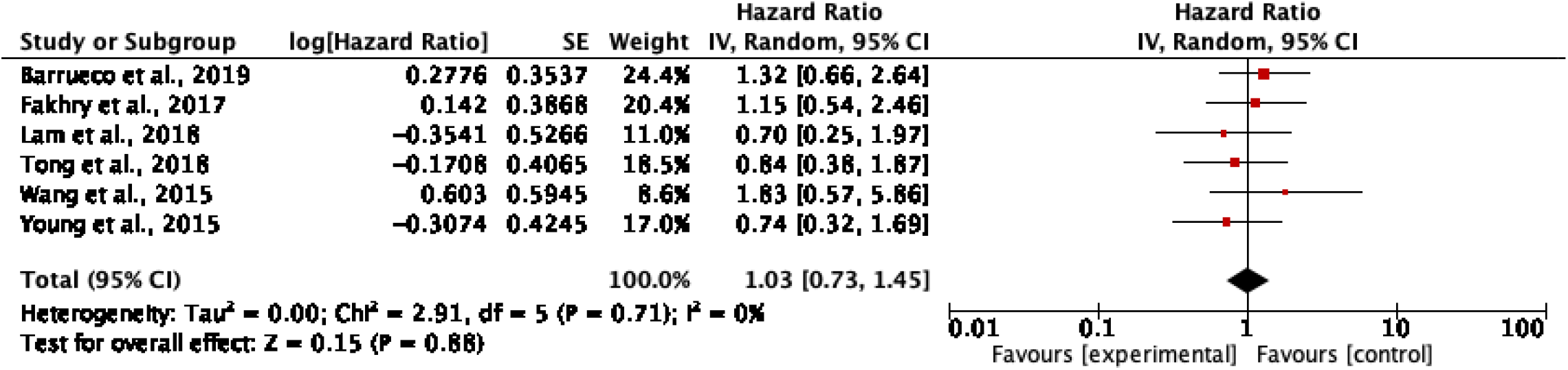
Association between p16 status and OS in laryngeal carcinoma Values on the right of the forest plot indicate reduced survival with p16 positivity. Meanwhile, values on the left of the plot indicate reduced survival with p16 negativity. The diamond represents the pooled result for all studies included in the analysis.

### HPV status and OS in laryngeal cancer

The data from seven studies was pooled for the meta-analysis on HPV status and OS in laryngeal cancer. This showed that there was an association between HPV status and OS in laryngeal cancer with an HR of 0.63 (95% CI: 0.41-0.97; p = 0.03) (S6). The test for heterogeneity showed moderate heterogeneity with an I^2^ of 57% and p-value of 0.03. The p value for Egger’s test indicated that there was no publication bias for OS (p = 0.562) (See Supplemental Materials). A Duval’s “Trim and Fill” analysis was performed, which suggested an additional two studies remain unpublished.

### QUIPS results

The Quality in Prognostic studies scoring suggested that most studies were low to moderate risk of bias (See Supplemental Materials). In regard to study participation, most studies were at low to moderate risk of bias, with one study at high risk of bias. For study attrition, again, most studies were at low to medium risk of bias, with one at high risk. This study was considered high risk due to failing to describe the reasons for patient inclusion/exclusion as well as failing to describe how many patients were excluded (29). For prognostic factor measurement, most were low to moderate risk of bias, with one at high risk of bias. The one study was high risk due to the authors not being able to determine the type of testing used for HPV status, as this varied across institutions and agencies included in the cohort (34). For outcome measurement, all studies were low to moderate risk of bias. For study confounding, the majority of studies were low to moderate risk of bias, with one at high risk of bias. This study was ranked as high risk for confounding due to the limited number of potential confounders measured (34). For statistical analysis and reporting, all 13 studies were found to be low risk of bias.

## Discussion

This meta-analysis sought to assess the prognostic value of HPV and p16 status on overall survival in LC and HPC. The pooled data demonstrated that p16 status was not significantly associated with OS in either LC or HPC with HRs of 1.03 (95% CI: 0.73, 1.45; p-value of 0.88) and 1.02 (95% CI: 0.55, 1.86, p-value = 0.96), respectively. Our results suggest that, unlike for OPSCC, p16 status does not have a significant impact on survival for LC or for HPC.

The pooled data did, however, reveal that HPV status was predictive of OS in LC with HR of 0.63 (95% CI: 0.41-0.97; p-value = 0.03). We did not pool the results for OS of HPC because only two studies reported it, nor did we pool the results for DFS or PFS for HPC due to only one study reporting each. This finding contradicts the current literature on HPV, which has suggested that the role of HPV on OS in LC is limited (40). However, the analysis was burdened by moderate heterogeneity. This may in part be due to the fact that the seven studies that were included in this study to assess prognosis for HPV in LC exhibited differences with respect to the proportion of HPV-positive tumors that were also p16-positive (28-30, 32, 34, 37, 38). For instance, Wang et. al found 14.28% of HPV DNA positive LCs were also p16 positive, whereas Tong et al. found that 84.8% of HPV DNA positive LCs were also p16 positive (28, 29). The other studies included in the analysis did not report the rate of concordance between HPV and p16 at all. This could be significant as previous literature in carcinoma of the tonsil and base of tongue has illustrated that a survival benefit exists when tumors are concurrently positive for p16 and HPV (41). It could also be due to the fact that HPV expression has been shown to vary significantly across LC and HPC. For instance, HPV DNA prevalence has been shown to be between 0 and 63% in LC and between 0 and 58% in HPC (14, 29, 38, 42, 43). In addition, significant variance in HPV prevalence across geographic areas has been previously noted and could also account for some of the heterogeneity (44). Thus, variation between the studies could also be explained by variation in HPV expression within the cohorts for each study.

It is important to note that although p16 and HPV status are found to correlate significantly in oropharyngeal cancer, current literature indicates that p16 status may not be as highly correlated with HPV status in non-oropharyngeal cancer (9, 16, 18). This suggests that there may be another mechanism besides HPV that is resulting in overexpression of p16 in these cancers. For instance, Chung et al. describe other tumor alterations, such as molecular aberration of the RB1 gene and chromosomal derangements, that could explain a rise in p16 tumor suppressor independent of HPV infection (9).

Our results highlight numerous opportunities for further research. The value of HPV status as a prognostic indicator in different treatment subgroups should be further studied, as differences between treatment subgroups have been noted in some of the literature. For instance, Tong et al. found that, although HPV status was not a significant prognostic indicator overall in laryngeal and hypopharyngeal cancer, HPV status was a positive prognostic indicator in the radiotherapy group (29, 45).

In addition, further research should be conducted to determine the optimal method of detecting p16 levels, as there is considerable variation in p16 expression patterns in LC and HPC reported in the published literature, which could be due to different detection methods and interpretation of results (15, 31, 46, 47). Finally, given the lack of conclusive evidence on the prognostic significance of HPV status for patients with LC or HPC, it will be important to investigate other potential prognostic factors with more clinical utility.

A limitation of this meta-analysis is that the majority of the studies included are retrospective cohort studies, which could place them at greater risk for bias in data collection and patient recruitment. Furthermore, it is important to interpret the results in the context of all-cause mortality, as most included studies do not differentiate between mortality due to HNC and mortality due to other causes. An advantage of this study is the low to moderate bias detected across most of the included studies. Only two studies were measured as high bias, each across two domains. Another advantage of the study is that there was no publication bias detected when analyzed via Begg’s funnel plot or Egger’s regression.

Overall, while the prognostic role of HPV and p16 is clearly defined in oropharyngeal cancers, there has been a growing body of contradictory research for LC and HPC (8-10, 12, 13). Our findings suggest that p16 is not associated with OS in LC and HPC. We are unable to rule out or interpret the role of HPV DNA in laryngeal carcinoma due to the high heterogeneity uncovered in our models. Overall, there is still insufficient published literature to determine definitively whether HPV status is associated with OS in LC and HPC. Physicians should use caution when considering HPV status when treating patients with LC or HPC.

## Data Availability

Data sharing not applicable. No new data generated.

## Acknowledgments

No acknowledgments to report for this paper.

## Notes

### Competing Interest Statement

The authors have declared no competing interest.

### Funding Statement

No external funding received for this project.

### Author Declarations

This was a systematic review and meta-analysis so registration with our Institutional Review Board (IRB) was not required.

