## Supplemental Tables and Figures for "The Impact of HPV DNA/p16 in Laryngeal/Hypopharyngeal Cancer: a Systematic Review and Meta-analysis"

**SUPPLEMENTAL DATA**

**S1: Search Strategy**

|  | ***Medline*** | ***Scopus*** | ***Embase*** | ***Cochrane*** |
| --- | --- | --- | --- | --- |
| Date Ran | 2.26.19 | 2.26.19 | 2.26.19 | 2.22.19 |
| Database Name | Pubmed-Medline | N/A | https://www.embase.com/#search | N/A |
| Search Limits | *((“cancer” OR “carcinoma” OR "Carcinoma"[Mesh] OR "carcinomas" OR “malignancies” OR “malignancy” OR “malignant” OR “neoplasia” OR “neoplasias” OR “neoplasm” OR “neoplasms” OR "Neoplasms"[Mesh])* ***AND***  ("Hypopharyngeal Neoplasms"[Mesh] OR "Laryngeal Neoplasms"[Mesh] *OR “laryngeal” OR “larynx” OR “laryngeal neoplasms” OR “laryngeal neoplasm” OR “laryngeal carcinoma” OR “laryngeal carcinomas” OR “laryngeal cancer” OR “laryngeal cancers” OR “carcinoma of the larynx” OR hypopharyn* OR “hypopharyngeal neoplasms” OR “hypopharyngeal neoplasm” OR “hypopharyngeal carcinoma” OR “hypopharyngeal carcinomas” OR “hypopharyngeal cancer” OR “hypopharyngeal cancers” OR “carcinoma of the hypopharynx”) AND ("Papillomaviridae"[Mesh] OR “papillomaviridae” OR “HPV”[tiab] OR “human papillomavirus” OR “human papillomaviruses” OR “human papilloma virus” OR “human papilloma viruses” OR “p16”[tiab] OR "Cyclin-Dependent Kinase Inhibitor p16"[Mesh]))* | ( LIMIT-TO ( DOCTYPE ,  "article" ) )  AND  ( LIMIT-TO ( EXACTKEYWORD ,  "Human" ) )  ***AND***   ( LIMIT-TO ( LANGUAGE ,  "English" ) )  AND  ( LIMIT-TO ( SRCTYPE ,  "journal" ) )  TITLE-ABS-KEY*(“cancer” OR “carcinoma” OR "carcinomas" OR “malignancies” OR “malignancy” OR “malignant” OR “neoplasia” OR “neoplasias” OR “neoplasm” OR “neoplasms”)*  ***AND***  TITLE-ABS-KEY(*“laryngeal” OR “larynx” OR “laryngeal neoplasms” OR “laryngeal neoplasm” OR “laryngeal carcinoma” OR “laryngeal carcinomas” OR “laryngeal cancer” OR “laryngeal cancers” OR “carcinoma of the larynx” OR hypopharyn* OR “hypopharyngeal neoplasms” OR “hypopharyngeal neoplasm” OR “hypopharyngeal carcinoma” OR “hypopharyngeal carcinomas” OR “hypopharyngeal cancer” OR “hypopharyngeal cancers” OR “carcinoma of the hypopharynx”)*  ***AND***  TITLE-ABS-KEY*(“papillomaviridae” OR “HPV” OR “human papillomavirus” OR “human papillomaviruses” OR “human papilloma virus” OR “human papilloma viruses” OR “p16” OR "Cyclin-Dependent Kinase Inhibitor p16")* | *((‘cancer’ OR ‘carcinoma’ OR ‘carcinomas’ OR ‘malignancies’ OR ‘malignancy’ OR ‘malignant’ OR ‘neoplasia’ OR ‘neoplasias’ OR ‘neoplasm’ OR ‘neoplasms’ OR 'neoplasm'/exp OR 'malignant neoplasm'/exp OR 'carcinoma'/exp OR 'malignancy'/exp)*  ***AND***  *(‘laryngeal cancer':ti,ab,kw OR ‘laryngeal neoplasms’*:ti,ab,kw *OR ‘laryngeal neoplasm’*:ti,ab,kw *OR ‘laryngeal carcinoma’*:ti,ab,kw OR ‘laryngeal cancer’:ti,ab,kw *OR 'hypopharyngeal cancer’*:ti,ab,kw*)*  ***AND*** *('Papillomaviridae'/exp OR ‘papillomaviridae’ OR ‘HPV’ OR ‘human papillomavirus’ OR ‘human papillomaviruses’ OR ‘human papilloma virus’ OR ‘human papilloma viruses’ OR 'Wart virus'/exp OR ‘p16’ OR ‘Cyclin-Dependent Kinase Inhibitor p16’ OR 'cyclin dependent kinase inhibitor 2A'/exp))* | ***#1***  MeSH descriptor: [Laryngeal Neoplasms] explode all trees  MeSH  288  ***#2***  MeSH descriptor: [Papillomaviridae] explode all trees  MeSH  541  ***#3*** (laryngeal):ti,ab,kw  ***AND*** (hpv):ti,ab,kw |

**S2: Summary of Screened Studies by Cancer Subtype, Country of Origin, Cohort Years, and Year of Publication**

| **Authors** | **Country of Origin** | **Cohort Years** | **Study Design** | **Subsite (HPC/LC)** | **Sample Size** | **Publication Year** |
| --- | --- | --- | --- | --- | --- | --- |
| Wang et al. | China | 1995-2009 | RCS | LC | 318 | 2015 |
| Tong et al. | China | N/A | RCS | LC | 211 | 2018 |
| Barrueco et al. | Spain | 1977-2005 | RCS | LC | 123 | 2019 |
| Young et al. | Australia | 2002-2012 | RCS | LC | 307 | 2015 |
| Yang et al. | China | 2000-2009 | RCS | LC | 196 | 2016 |
| Wendt et al. | Sweden | 2000-2007 | RCS | HPC | 109 | 2014 |
| Meshman et al. | USA | 2009-2014 | RCS | LC & HPC | 31 | 2017 |
| Shaughnessy et al. | USA | 2009-2011 | RCS | LC & HPC | 47 | 2014 |
| Lopez et. al | Brazil | 1998-2008 | RCS | LC | 398 | 2014 |
| Lam et al. | China | 2005-2010 | RCS | LC | 85 | 2018 |
| Joo et al. | South Korea | 2004-2011 | RCS | HPC | 45 | 2014 |
| Joo et al. | South Korea | 1996-2011 | RCS | HPC | 64 | 2013 |
| Dahm et al. | Austria | 2007-2016 | RCS | LC & HPC | 134 | 2018 |
| Chung et al. | N/A | N/A | RCS | LC & HPC | 683 | 2014 |
| Li et al. | USA | 2010-2014 | RCS | LC & HPC | 9656 | 2018 |
| Lee et al. | South Korea | 2004-2013 | RCS | HPC | 45 | 2018 |
| Ang et al. | Singapore | 2009-2011 | RCS | HPC | 75 | 2014 |
| Fakry et al. | USA | 1995-2012 | RCS | LC | 243 | 2017 |
| Pietruszewska et al. | Poland | NR | RCS | LC | NR | 2007 |
| Wang et al. | China | NR | RCS | LC | 186 | 2014 |
| Vlachtsis et al. | N/R | 1999-2002 | RCS | LC | 212 | 2005 |
| Tiefenbock-Hansson et al. | Sweden | 1999-2010 | RCS | LC | 149 | 2017 |
| Tehrany et al. | Germany | 1992-2011 | RCS | All HNSCC | 233 | 2015 |
| Stephen et al. | USA | 1986-2003 | RCS | All HNSCC | 80 | 2013 |
| Stephen et al. | USA | NR | RCS | LC | 79 | 2012 |
| Smith et al. | USA | 1994-2004 | RCS | LC & HPC | 294 | 2008 |
| Sivars et al. | Sweden | 2000-2007 | Review | HPC | NR | 2016 |
| Silva et al. | Brazil | NR | RCS | LC | 35 | 2012 |
| Sanchez Barruecho et al. | Spain | 1977-2005 | RCS | LC | 123 | 2017 |
| Salazar et al. | USA | NR | RCS | All HNSCC | 222 | 2014 |
| Salazar et al. | USA | NR | RCS | LC & HPC | 58 | 2014 |
| Rivera-Pena et al. | Puerto Rico | 1993-2005 | RCS | LC | 185 | 2016 |
| Riaz et al. | USA | 2006-2008 | RCS | LC & HPC | 17 | 2016 |
| Reka Fejer et al. | Hungary | NR | RCS | All HNSCC | 81 | 2016 |
| Rautava et al. | Canada | 1988-2009 | RCS | LC & HPC | 25 | 2012 |
| Rades et al. | Germany | NR | RCS | LC & HPC | 24 | 2011 |
| Zhao et al. | USA | 2002-2006 | RCS | All HNSCC | 143 | 2012 |
| Zackrisson et al. | Sweden | 1998-2006 | RCS | All HNSCC | 750 | 2015 |
| Yang et al. | China | 2001-2008 | RCS | LC | 38 | 2018 |
| **Authors** | **Country of Origin** | **Cohort Years** | **Study Design** | **Subsite (HPC/LC)** | **Sample Size** | **Publication Year** |
| Xu et al. | China | 2006-2013 | LCS | LC | 674 | 2014 |
| Xiao et al. | USA | 2002-2005 | RTOG Trial | All HNSCC | 743 | 2017 |
| Wilson et al. | USA | 2002-2011 | Chart Review | HPC | 27 | 2012 |
| Wilson et al. | USA | 2002-2013 | Chart Review | HPC | 32 | 2014 |
| Wildeman et al. | The Netherlands | 1985-1999 | Case-Control Study | LC | 59 | 2009 |
| Pintos et al. | Canada | 1982-1992 | RCS | LC | 44 | 1999 |
| Picard et al. | France | 1994-2014 | Retrospective Monocentric Study | LC & HPC | 15 | 2016 |
| Peralta et al. | Mexico | 2004-2008 | RCS | LC | 30 | 2018 |
| Lundberg et al. | Finland | 1997-2008 | RCS | LC & HPC | 130 | 2016 |
| Lohaus et al. | Germany | 2005-2010 | RCS | HPC | 35 | 2014 |
| Lassen et al. | Denmark | 1970-2012 | RCS | LC & HPC | 479 | 2014 |
| Karpathiou et al. | Australia | NR | RCS | LC & HPC | 68 | 2016 |
| Kanyilmaz et al. | Turkey | 2006-2010 | RCS | LC & HPC | 112 | 2015 |
| Kalfert et al. | Czech Republic | 2001-2009 | RCS | LC | 58 | 2014 |
| Jiang et al. | China | 1997-2008 | RCS | LC | 71 | 2013 |
| Hoffmann et al. | Germany | 1994-2002 | RCS | LC & HPC | 43 | 2005 |
| Hernandez et al. | USA | 1993-2004 | RCS | LC | 148 | 2016 |
| Deng et al. | Japan | 2006-2013 | PCS | LC & HPC | 63 | 2014 |
| Davidson et al. | USA | 2010-2012 | ROCS | LC | 3238 | 2018 |
| Dalianis et al. | Sweden | 2008-2013 | RCS | HPC | 93 | 2015 |
| Clayman et al. | USA | NR | Chart Review | LC & HPC | 78 | 1994 |
| Cho et al. | North Korea | NR | Chart Review | LC & HPC | 15 | 2017 |
| Chen et al. | Taiwan | 2006-2009 | RCS | LC | 106 | 2017 |
| Burr et al. | USA | 2010-2013 | RCS | LC & HPC | 293 | 2018 |
| Bryant et al. | USA | 2005-2015 | RCS | LC & HPC | 259 | 2018 |
| Brewczynski et al. | Egypt | 2009-2014 | RCS | LC & HPC | 168 | 2017 |
| Brandwein et al. | USA | 1988-1991 | RCS | LC | 40 | 1993 |
| Boelke et al. | Germany | NR | RCS | LC & HPC | 28 | 2017 |
| Birtalan et al. | Hungary | NR | RCS | LC & HPC | 65 | 2018 |
| Biesaga et al. | Poland | 2007-2014 | RCS | LC & HPC | 17 | 2018 |
| Bentzen et al. | Denmark | NR | Randomized trial | LC & HPC | 54 | 2015 |
| Baez et al. | Puerto Rico | NR | RCS | LC | 52 | 2004 |
| Badaracco et al. | Italy | NR | RCS | LC & HPC | 35 | 2007 |
| Atighechi et al. | Iran | 2007-2012 | Case-control study | LC | 44 | 2016 |
| Asvadi-Kermani et al. | Iran | 2008-2010 | Case-control study | LC & HPC | 7 | 2012 |
| **Authors** | **Country of Origin** | **Cohort Years** | **Study Design** | **Subsite (HPC/LC)** | **Sample Size** | **Publication Year** |
| Antonsson et al. | Australia | 2004-2010 | Case-control study | LC & HPC | 83 | 2015 |
| Allegra et al. | Italy | 2009-2010 | Case-control study | LC | 31 | 2013 |
| Habbous et al. | Canada | 2000-2010 | RCS | LC & HPC | 1640 | 2014 |
| Gudlvicience et al. | Lithuania | 2006 | Case-control study | LC & HPC | 20 | 2012 |
| Flores de-la Torre et al. | Mexico | 2003-2007 | RCS | LC | 59 | 2010 |
| Fakhry et al. | USA | NR | PCS | LC | 34 | 2008 |
| Ernoux-Neufcoeur et al. | France | 1996-2000 | N/A | HPC | 75 | 2011 |
| Erkul et al. | Turkey | 2005-2014 | N/A | LC | 78 | 2017 |
| Duray et al. | Belgium | NR | N/A | LC & HPC | 24 | 2013 |
| Du et al. | USA | 2002-2006 | PCS | LC & HPC | 306 | 2019 |
| Donmez et al. | Not English | Not English | Not English | Not English | Not English | 2012 |
| Adkins et al. | USA | NR | Clinical Trial | LC & HPC | 12 | 2016 |
| Acuna et al. | Spain | 2000-2017 | Case-control study | LC & HPC | 10 | 2018 |

*RCS, retrospective cohort study; PCS, prospective cohort study; ROCS, retrospective observational cohort study; LCS, longitudinal cohort study.*

**S3: Summary of Excluded Full-Text Articles (with reasons)**

| **Authors** | **Year** | **Reason** |
| --- | --- | --- |
| Pietruszewska et al. | 2007 | Not English |
| Wang et al. | 2014 | Not English |
| Vlachtsis et al. | 2005 | No HR Reported |
| Tiefenbock-Hansson et al. | 2017 | No HR Reported |
| Tehrany et al. | 2015 | No HR Reported |
| Stephen et al. | 2013 | No HR Reported |
| Stephen et al. | 2012 | No Survival Data |
| Smith et al. | 2008 | No HR Reported |
| Sivars et al. | 2016 | Review Paper |
| Silva et al. | 2012 | No HR Reported |
| Sanchez Barruecho et al. | 2017 | No HR Reported |
| Salazar et al. | 2014 | No HR Reported |
| Salazar et al. | 2014 | No HR Reported |
| Rivera-Pena et al. | 2016 | No HR Reported |
| Riaz et al. | 2016 | No HR Reported |
| Reka Fejer et al. | 2016 | No HR Reported |
| Rautava et al. | 2012 | No HR Reported |
| Rades et al. | 2011 | No HR Reported |
| Zhao et al. | 2012 | p16 Localization Study |
| Zackrisson et al. | 2015 | No HR Reported |
| Yang et al. | 2018 | No HR Reported |
| Xu et al. | 2014 | No HR Reported |
| Xiao et al. | 2017 | No HR Reported |
| Wilson et al. | 2012 | No HR Reported |
| Wilson et al. | 2014 | No HR Reported |
| Wildeman et al. | 2009 | No HR Reported |
| Pintos et al. | 1999 | No HR Reported |
| Picard et al. | 2016 | No HR Reported |
| Peralta et al. | 2018 | No HR Reported |
| Lundberg et al. | 2016 | No HR Reported |
| Lohaus et al. | 2014 | No HR Reported |
| Lassen et al. | 2014 | No HR Reported |
| Karpathiou et al. | 2016 | No HR Reported |
| Kanyilmaz et al. | 2015 | No HR Reported |
| Kalfert et al. | 2014 | No HR Reported |
| Jiang et al. | 2013 | No HR Reported |
| Hoffmann et al. | 2005 | No HR Reported |
| Deng et al. | 2014 | No HR Reported |
| Davidson et al. | 2018 | No HR Reported |
| Dalianis et al. | 2015 | No HR Reported |
| Clayman et al. | 1994 | No HR Reported |
| Cho et al. | 2017 | Wrong Histological Subtype |
| Chen et al. | 2017 | No HR Reported |
| Burr et al. | 2018 | No HR Reported |
| Bryant et al. | 2018 | No HR Reported |
| Brewczynski et al. | 2017 | Abstract Only |
| Brandwein et al. | 1993 | Prevalence Analysis |
| Boelke et al. | 2017 | Abstract Only |
| Birtalan et al. | 2018 | No HR Reported |
| Biesaga et al. | 2018 | No HR Reported |
| **Authors** | **Year** | **Reason** |
| Bentzen et al. | 2015 | No HR Reported |
| Baez et al. | 2004 | No HR Reported |
| Badaracco et al. | 2007 | No HR Reported |
| Atighechi et al. | 2016 | No HR Reported |
| Asvadi-Kermani et al. | 2012 | No HR Reported |
| Antonsson et al. | 2015 | No HR Reported |
| Allegra et al. | 2013 | No HR Reported |
| Habbous et al. | 2014 | No HR Reported |
| Gudlvicience et al. | 2012 | No HR Reported |
| Flores de-la Torre et al. | 2010 | Wrong Histological Subtype |
| Fakhry et al. | 2008 | No HR Reported |
| Ernoux-Neufcoeur et al. | 2011 | No HR Reported |
| Erkul et al. | 2017 | No HR Reported |
| Duray et al. | 2013 | No HR Reported |
| Du et al. | 2019 | No HR Reported |
| Donmez et al. | 2012 | Not in English |
| Adkins et al. | 2016 | No HR Reported |
| Acuna et al. | 2018 | Wrong Histological Subtype |

**S4. Quality in Prognostic Studies (QUIPS) Criteria Scoring**

|  | Low | Medium | High |
| --- | --- | --- | --- |
| Study Participation | 9 | 4 | 1 |
| Study Attrition | 10 | 3 | 1 |
| Prognostic Factor Measurement | 8 | 5 | 1 |
| Outcome Measurement | 12 | 2 | 0 |
| Study Confounding | 12 | 1 | 1 |
| Statistical Analysis and Reporting | 14 | 0 | 0 |

*Figure graphically depicts the risk of bias assessment conducted using the Quality In Prognosis Studies tool (QUIPS).The y-axis represents the percentage of studies graded to a specific risk of bias: low, moderate, or high risk of bias. The x-axis represents the 6 domains that were graded: study participation, study attrition, prognostic factor measurement, outcome assessment, confounding factors, and statistical analysis and reporting.*

**S5. Duval and Tweedie’s Trim and Fill Plot for p16 and OS in Laryngeal Cancer**

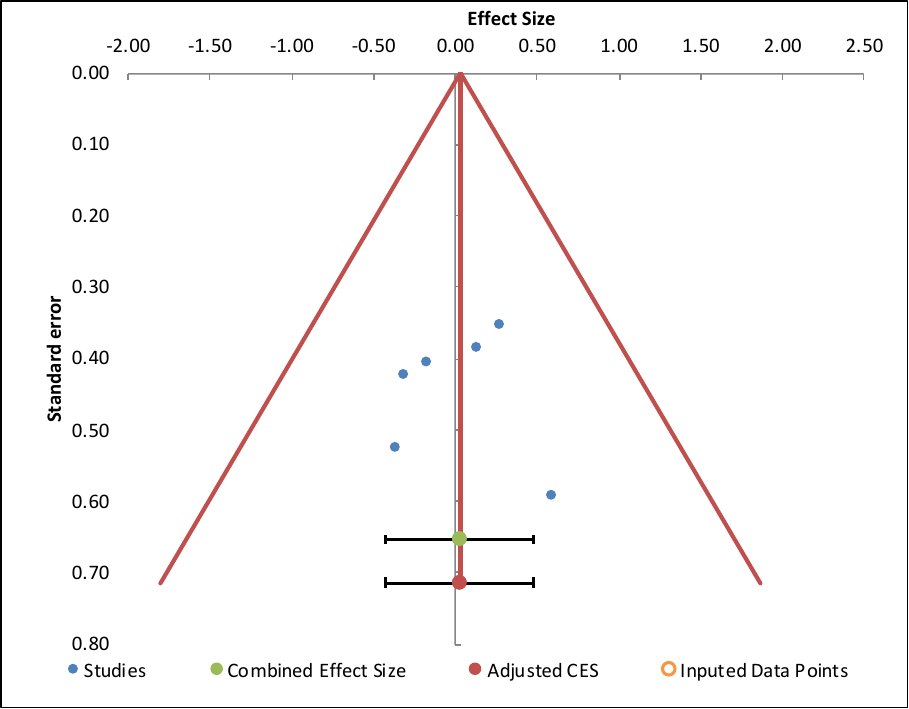

| **Combined effect size** | **Observed** |
| --- | --- |
| Effect Size | 0.03 |
| SE | 0.17 |
| CI Lower limit | -0.42 |
| CI Upper limit | 0.48 |
| PI Lower limit | -0.42 |
| PI Upper limit | 0.48 |
| **Combined effect size** | **Adjusted** |
| Effect Size | 0.03 |
| SE | 0.17 |
| CI Lower limit | -0.42 |
| CI Upper limit | 0.48 |
| PI Lower limit | -0.42 |
| PI Upper limit | 0.48 |

| **Egger Regression** | | | | |
| --- | --- | --- | --- | --- |
|  | Estimate | SE | CI LL | CI UL |
| Intercept | -0.07 | 2.06 | -5.38 | 5.23 |
| Slope | 0.06 | 0.88 | -2.22 | 2.33 |
| t test | -0.04 |  |  |  |
| p-value | 0.973 |  |  |  |

**Failsafe tests**

| Overall Z-score | 0.14 |
| --- | --- |
| Failsafe-N | 0 |
| Ad-hoc rule | TRUE |

**S6. Association between HPV status and OS in laryngeal cancer**

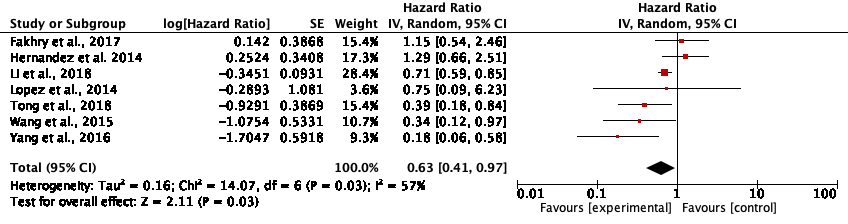

Values on the right of the forest plot indicate reduced survival with HPV-positivity. Meanwhile, values on the left of the plot indicate reduced survival with HPV negativity. The diamond represents the pooled result for all studies included in the analysis.

**S7. Duval and Tweedie’s Trim and Fill Plot for HPV and OS in Laryngeal Cancer**

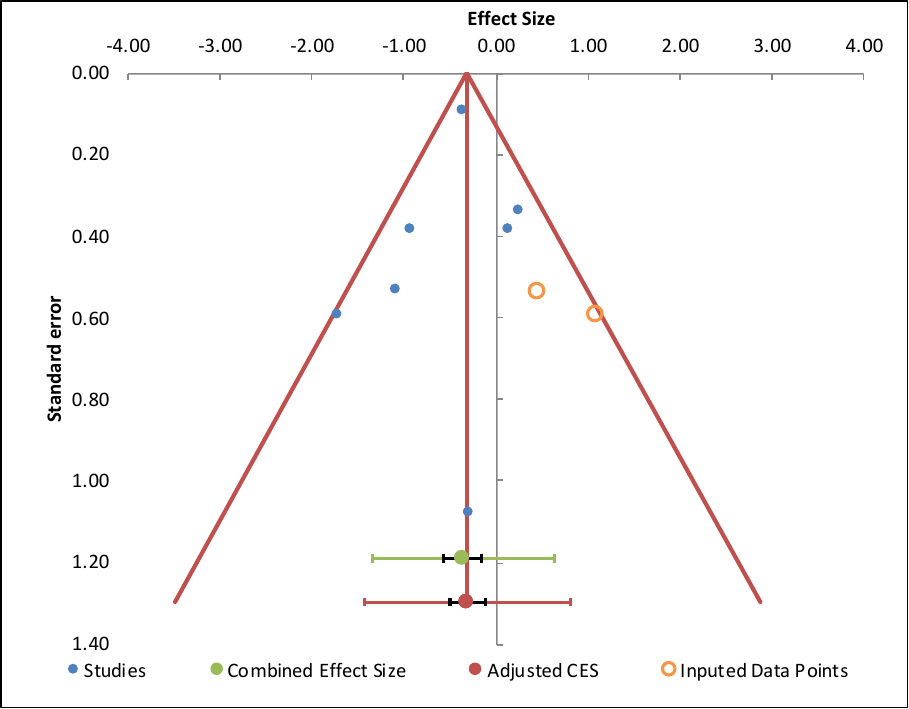

| **Combined effect size** | **Observed** |
| --- | --- |
| Effect Size | -0.36 |
| SE | 0.08 |
| CI Lower limit | -0.56 |
| CI Upper limit | -0.15 |
| PI Lower limit | -1.35 |
| PI Upper limit | 0.63 |
| **Combined effect size** | **Adjusted** |
| Effect Size | -0.31 |
| SE | 0.08 |
| CI Lower limit | -0.50 |
| CI Upper limit | -0.12 |
| PI Lower limit | -1.43 |
| PI Upper limit | 0.80 |

| **Egger Regression** | | | | |
| --- | --- | --- | --- | --- |
|  | Estimate | SE | CI LL | CI UL |
| Intercept | -0.56 | 0.90 | -2.76 | 1.64 |
| Slope | -0.27 | 0.20 | -0.75 | 0.22 |
| t test | -0.62 |  |  |  |
| p-value | 0.562 |  |  |  |

| **Failsafe tests** | |
| --- | --- |
| Overall Z-score | -3.84 |
| Failsafe-N | 31 |
| Ad-hoc rule | TRUE |
